# IMPROVE-DD: Integrating Multiple Phenotype Resources Optimises Variant Evaluation in genetically determined Developmental Disorders

**DOI:** 10.1101/2022.05.20.22275135

**Authors:** Stuart Aitken, Helen V Firth, Caroline F Wright, Matthew E Hurles, David R FitzPatrick, Colin A. Semple

## Abstract

**Summary:** Diagnosing rare developmental disorders using genome-wide sequencing data commonly necessitates review of multiple plausible candidate variants, often using ontologies of categorical clinical terms. We show that Integrating Multiple Phenotype Resources Optimises Variant Evaluation in Developmental Disorders (IMPROVE-DD) by incorporating additional classes of data commonly available to clinicians and recorded in health records. In doing so, we quantify the distinct contributions of gender, growth, and development in addition to Human Phenotype Ontology (HPO) terms, and demonstrate added value from these readily-available information sources. We use likelihood ratios for nominal and quantitative data and propose a novel classifier for HPO terms in this framework. This Bayesian framework results in more robust diagnoses. Using data systematically collected in the DDD study, we considered 77 genes with pathogenic/likely pathogenic variants in ≥10 probands. All genes showed at least a satisfactory prediction by ROC when testing on training data (AUC≥0.6), and HPO terms were the best individual predictor for the majority of genes, though a minority (13/77) of genes were better predicted by other phenotypic data types. Overall, classifiers based upon multiple integrated phenotypic data sources performed better than those based upon any individual source, and importantly, integrated models produced notably fewer false positives. Finally, we show that IMPROVE-DD models with good predictive performance on cross-validation can be constructed from relatively few cases. This suggests new strategies for candidate gene prioritisation, and highlights the value of systematic clinical data collection to support diagnostic programmes.

---

The importance of phenotype to ranking candidate disease-causing genes is established in research and increasingly so in clinical practice. The primary data resource used in computational phenotype analyses is the Human Phenotype Ontology (HPO)^1^. Despite promising results^2,3^, the exploitation of other information readily available to clinicians, including quantitative anatomic measurements and patient images, is less prevalent^4^. The HPO resource, HPO-encoded disease models and patient’s disease descriptions in HPO terms support diverse tasks: protocols start with a systematic description of an individual’s phenotype and may progress to suggested diagnoses^5^. While numerous computational and statistical approaches have been proposed, the advantages of *likelihood ratios* for the interpretation of genomic and phenomic data in rare disease have been demonstrated^6^.

Probabilistic methods for combining HPO terms with genetic data in Mendelian disease have beed proposed^7-9^, as have statistical criteria^10-12^ and deep learning^13,14^, commonly as component of a variant prioritisation workflow. Others have aimed to support users through ontology-assisted visualisation and ranking^15-18^. Here, we show that Integrating Multiple Phenotype Resource Optimises Variant Evaluation in Developmental Disorders (IMPROVE-DD) by utilising a range of clinical datasets coupled with gold-standard diagnoses confirmed by clinical evaluation.

Probabilistic models are often compared through the likelihood ratio which uses Bayes rule to decompose the joint probability of the models under consideration (*M*_1_ and *M*_*2*_) and the data to the conditional probability of the data given the model and the prior:

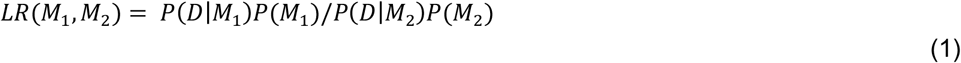

This formulation avoids calculating the probability of observing the data *P(D)* which can be difficult to evaluate.

The Deciphering Developmental Disorders (DDD) study recruited individuals with severe or extreme developmental disorders (DD) in whom clinical assessment and baseline genetic investigation were unable to establish a molecular diagnosis^19-22^. Whole exome sequencing (WES) was performed in >13,500 unrelated probands with 85% analysed as nuclear trios (affected child with both parents) and the remainder as singleton WES. Detailed phenotypic information (see below) was recorded by clinicians using the secure portal within the DECIPHER system^23^ (deciphergenomics.org). A combination of rational filtering for impactful genotypes at known DD loci and statistical genomic approaches to the discovery of novel loci and genetic mechanisms has proven to be successful in diagnosing in >33% of the cohort.

At the time of access, the DDD dataset included information on 13,439 probands. In addition to age, gender and HPO terms, the available clinical information included: five growth attributes: gestation (in months), birthweight, height, weight and head circumference (expressed as z-scores); plus four developmental milestones (in months) marking: when the child first walked independently, spoke their first words, expressed a social smile, and sat independently.

We used genetic diagnoses assigned by referring clinical centres as the ground truth and only considered cases with a single diagnosis. Diagnoses in one of 856 genes were recorded for 4,112 probands. However, we required at least 10 probands per genetic disorder to build gene-disease models, and a relatively complete record of quantitative data in order to be able to model a gene. This reduced the number of genes we could consider to 77 in 1,730 probands. The median number of probands per gene was 17 (maximum 81).

In IMPROVE-DD (Web Resources), we apply Bayesian methods to integrate diverse quantitative phenotypic data types and measure their contribution to decision making. Here, decision-making is formalised as classifying each case to the correct genetic diagnosis (for example, *ADNP* as the causal gene vs not), to test whether the phenotype under consideration is both consistently observed, and sufficiently distinct from that of the remaining DD cohort. We took advantage of the likelihood ratio approach to explore the contributions to making a diagnosis of each of the available data types available in DDD (gender, growth, development and HPO annotations) by implementing a naïve Bayes classifier for each gene from each data type (**Figure 1A** and **1B**) using the R package naivebayes (Web Resources). Each classifier computes a likelihood ratio (equation 1) comparing *M*_1_ *w*ith 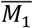 taking the prior as the observed frequency of each hypothesis in the data. A classifier for nominal data such as gender is simply a table of probabilities, while for continuous data we used a smoothed kernel (the nrd0 kernel with increased bandwidth) to model the data. A feature will have diagnostic value when its distribution for *M*_1_ *d*iffers from that of 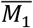, for example, for *ARID1B*, head circumference (OFC) is discriminative but weight is not (**Figure 1A**).

**Figure 1.**
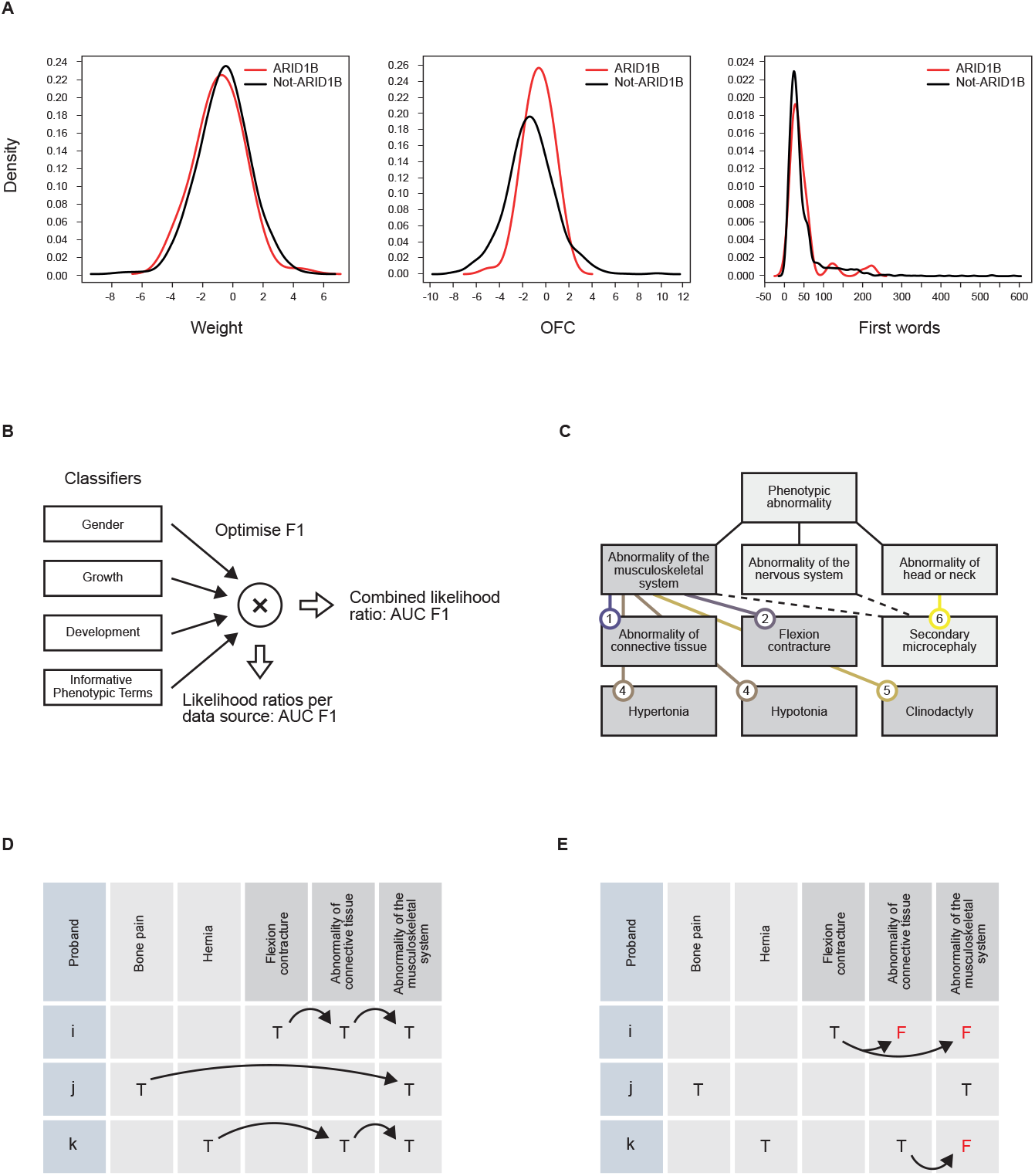
Integrating probabilistic models for nominal, continuous and ontology annotation data. (A) Continuous data for growth and development is modelled by a smoothed kernel density. (B) Classifiers for gender, growth, development and HPO terms are constructed and evaluated individually, and are combined by optimally weighting their output likelihoods. (C) All top-level HPO phenotypes (immediately below *Phenotypic abnormality*) are considered informative phenotypic terms (IPTs). Dependent on their annotation frequency across the cohort, child terms are selected as IPTs (250-1500 uses) or further expanded (>1500 uses). Duplicate child terms such as *Secondary microcephaly* are included under one top-level IPT only. The number of HPO levels a child term is below its top-level term is given at the end of the arc. (D) Annotations are propagated from child to parent terms (arrows) to expand the annotation matrix according to the HPO structure. (E) Annotations to IPTs that arise from propagation from an IPT are removed: Proband *i* has *Flexion contracture* but not parents of this term whereas proband *j* has *Abnormality of the musculoskeletal system* among the annotations to informative terms as *Bone pain* is not selected as informative.

We next sought methods for selecting HPO terms and deriving useful probabilistic models from them in the likelihood framework. Information content (IC), defined as -log(probability of term)^24^ has been combined with genomic freqency, and has been used to compare ontology-encoded phenotypes to aid prediction^8,20^. When considering the entire corpus of annotations, the least frequently used terms are most informative but describe the fewest cases. First, annotation frequencies for all terms, whether used directly in annotation or not, were found by propagating all annotations to their parent terms. A set of *informative phenotypic terms* (IPTs) was identified as follows: starting with the top-level terms for phenotypic abnormality that distinguish disorders of the major organ systems and developmental processes, each was expanded into a set of child terms that met minimal and maximal annotation frequency criteria across the entire cohort (2% and 10% respectively) (**Figure 1C**). Working in a top-down manner to preferentially select terms that balance IC with generality in the ontology graph, child terms with usage above the upper limit were expanded, those within lower and upper limits were retained in the IPT set. Duplicate IPTs were deleted to ensure annotations to IPTs were independent (**Figure 1D** and **1E**). The frequency of use of IPTs was found from the modified annotation matrix in a computationally efficient manner. Examples of informative phenotypic terms included Mild, Moderate and Severe global developmental delay (HP:0011342, HP:0011343, HP:0011344) organised under top-level IPT Abnormality of the nervous system (HP:0000707) (**Table 1, Table S1**).

**Table 1.** Examples of informative phenotypic terms.

| Abnormality of the nervous system (HP:0000707) | Abnormality of the cardiovascular system (HP:0001626) | Abnormality of limbs (HP:0040064) |
| --- | --- | --- |
| Mild global developmental delay (HP:0011342) | Abnormal heart morphology (HP:0001627) | Abnormal 5<sup>th</sup> finger morphology (HP:0004207) |
| Moderate global developmental delay (HP:0011343) | Abnormality of the vasculature (HP:0002597) | Abnormal thumb morphology (HP:0001172) |
| Severe global developmental delay (HP:0011344) | Abnormal cardiovascular system physiology (HP:0011025) | Abnormal fingertip morphology (HP:0001211) |

This procedure uses term frequencies from annotations to all probands irrespective of their diagnosis, seeking terms with moderate IC across the dataset. The resulting 157 terms were used as features in classifiers for all genes we modelled: their frequency of use was found from the modified annotation matrix for IPTs when it was factored by proband diagnosis (Supplemental Methods).

Measures from information retrieval including term frequency (TF) and inverse document frequency (IDF) have previously been adopted for the selection of relevant HPO terms^25^. To compare our set of IPTs with those from an information theoretic (TF IDF IC) approach, we computed these measures for our dataset and examined the position of our IPTs in a ranking of terms per gene (**Figure 2A**). IPTs seldom ranked highest by TF IDF IC, and can rank rather lowly, thus would not likely be selected by such a method. Terms ranking highly by TF IDF IC had considerably higher information content than IPTs, but considering the top 10 such terms, the number of terms per proband was low (**Figure 2B**). Applying the same ranking procedure to the terms that define a HPO gene model^1,26^(see Web Resources), we observed a more uniform ranking of HPO terms, some terms ranked highly and others very lowly (**Figure S1**). We conclude that our approach uncovers a set of terms that are unlikely to have been selected by conventional metrics.

**Figure 2.**
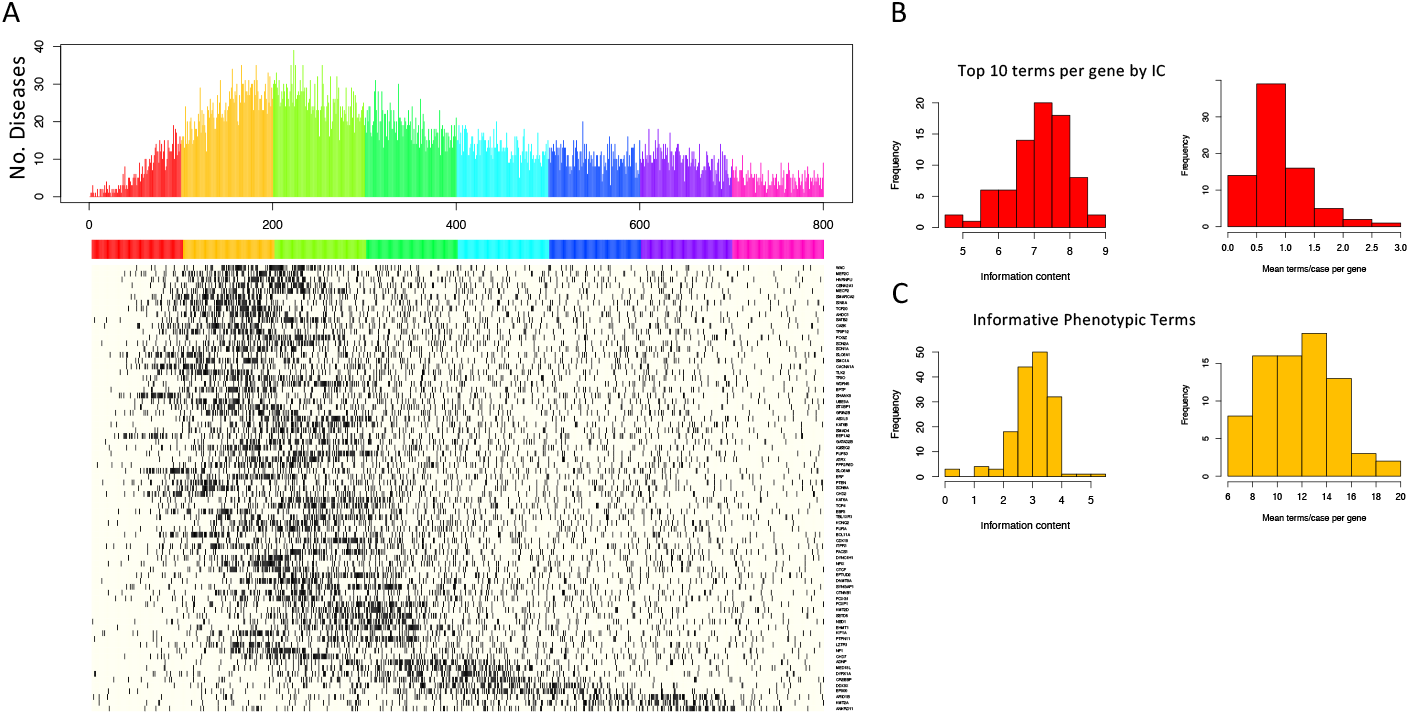
Informative phenotypic terms differ from the highest scoring terms by information theoretic criterion. Heatmap showing for each gene (row) the occurrence of an IPT in a ranking of HPO terms by TF IDF IC. HPO terms are ordered left to right by decreasing TF IDF IC. Top panel shows the number of diseases for which an informative term is found in rank *i* (from 1 to 800), colours indicate scale, each covers 100 positions. Although occurring towards the top of the 2634 terms for which this measure can be calculated, IPTs are seldom in the top 50, and for genes such as *ANKRD11, ARID1B* and *KMT2A* in the bottom rows rank 400 and below. (B) Histograms of mean term IC (left) and mean number of probands per term (right) when selecting the top 10 terms scored by TF IDF IC per gene. (C) Histograms of term IC (left) and number of probands per term (right) for the 157 IPTs.

As summary statistics we report the F1 score, being the harmonic mean of precision and recall, as a measure of the accuracy of decision making, and AUC as a measure of the rank of true positives irrespective of decision threshold across the entire dataset. The individual classifier outputs were combined by a set of weights, found per gene, that optimised F1 from the four input likelihoods plus an additional constant representing the prior. We discuss F1 and AUC from testing on the training data as indicative of the potential of phenotype modelling, and results from a leave-one-out cross-validation which better controls for model overfitting.

Beginning with the results from testing on the training data, the IPT-based HPO classifier had the best performance in decision making for most genes (**Figure 3A**) with F1 scores of up to 0.46 when testing on training data. Exceptions were apparent, for example, growth was a better predictor for *NSD1*, while development was a better predictor for *GRIN2B*. Genes with larger numbers of cases tended to score well when testing on training data: Pearson correlation between F1 from the HPO classifier and the number of probands per gene was 0.61 (0.51 in cross-validation; p<1e-5) indicating that performance was positively influenced by the number of cases. In the interpretation of F1 scores it should be noted that the prior probabilities *p*(*M*_0_) derived from the number of cases range from 1/150 to 1/19 whereas the alternative hypotheses are much more probable (95/100 to 99/100). Consequently, the evidence from the data commonly failed to outweigh the prior and hence precision or recall was 0 and F1 could not be calculated.

**Figure 3.**
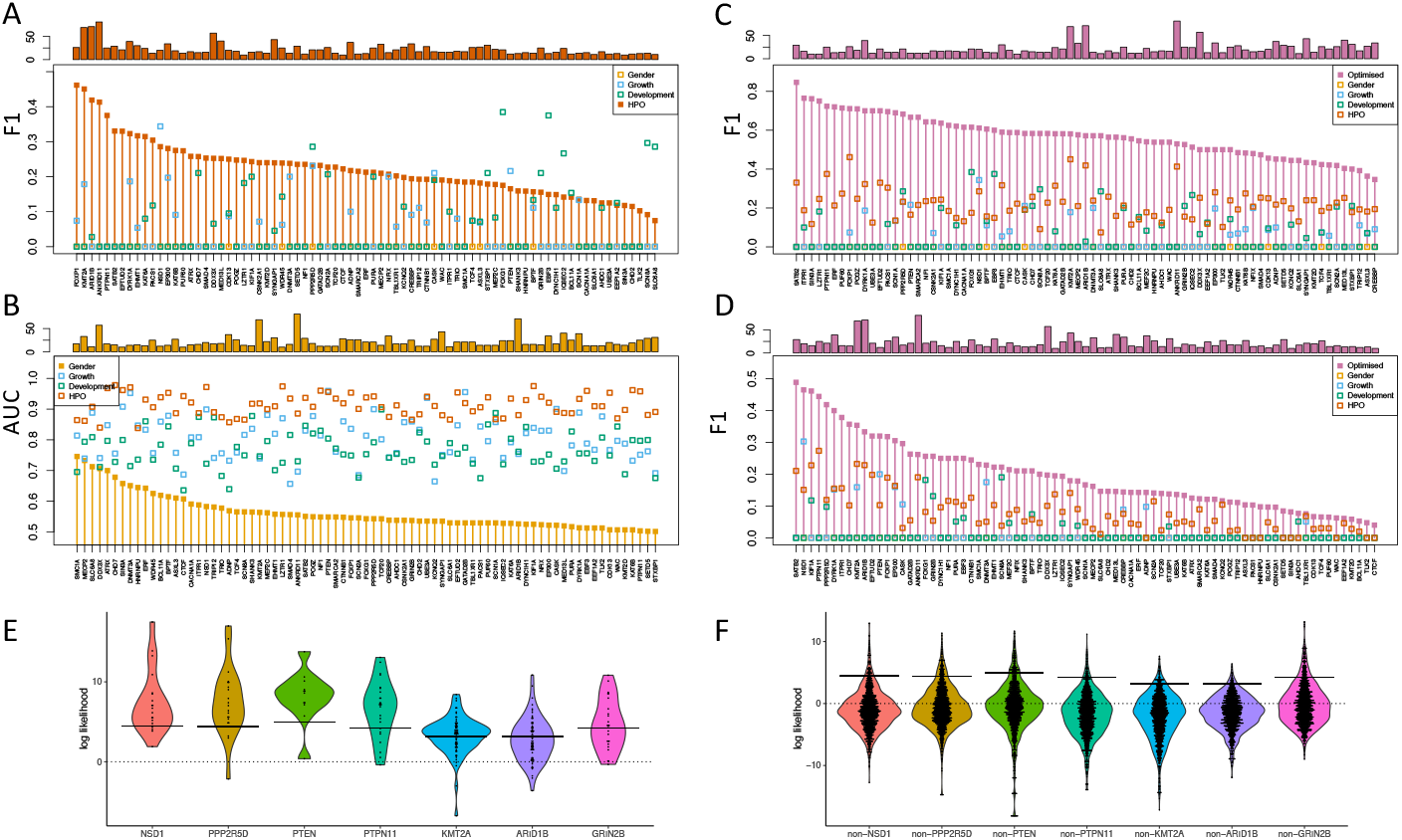
Integrating multiple phenotypic models improves classification. (A) F1 per gene from gender, growth, development and HPO classifiers (all values plotted at the same x coordinate). Vertical bars and filled symbols highlight the HPO classifier performance. Number of probands per gene (upper). (B) AUC per gene highlighting gender. (C) F1 per gene highlighting the optimised score. (D) F1 per gene highlighting the optimised score when combining likelihoods from cross-validation. (E) Violin plots of likelihood ratios from HPO terms for selected genes (symbols are probands). Likelihoods are shown for a prior of 0.5 to give a common zero reference line for all models. The per-gene prior is shown by the horizontal bar. The HPO model may represent a consensus for a majority of probands, yet the model may not be sufficient to give a diagnosis for some individuals (*KMT2A, ARID1B*). Alternatively, the phenotypic spectrum may be broad (*NSD1, PPP2R5D*) and correctly diagnose all but a few cases. (F) Violin plots for the gene models in (E) applied to probands whose diagnosis differs. A log likelihood above the per-gene prior is a false positive.

The best F1 scores from growth data alone were from *NSD1, PTEN* and *DNMT3A* (**Figure 3A, S2, S3**) with 20, 12 and 14 cases respectively (top three on cross-validation and top 5 when testing on training data). Turning to developmental milestones, the best F1 scores were from *SCN8A, FOXG1* and *GRIN2B* (**Figure 3A, S2, S3**) with 14, 21 and 25 cases respectively (top 3 by cross-validation and in the top 8 testing on training data).

The best predicted genes from HPO annotations were *PTPN11, KMT2A* and *ARID1B* with 25, 69 and 71 cases respectively (top 3 on leave-one-out cross-validation, top 5 when testing on training data). The likelihoods of individual IPTs can be examined for each proband and provide potentially useful diagnostic information to a clinician. We list the three most likely and three least likely *KMT2A* diagnoses according to phenotype modelling to show the balance of HPO terms for and against, and, in the final case, the positive contribution from growth outweighed by the negative contribution from HPO terms (**Table 2)**. Of note, the top three cases are pathogenic frameshifts, whereas the bottom three are likely pathogenic missense variants.

**Table 2.**
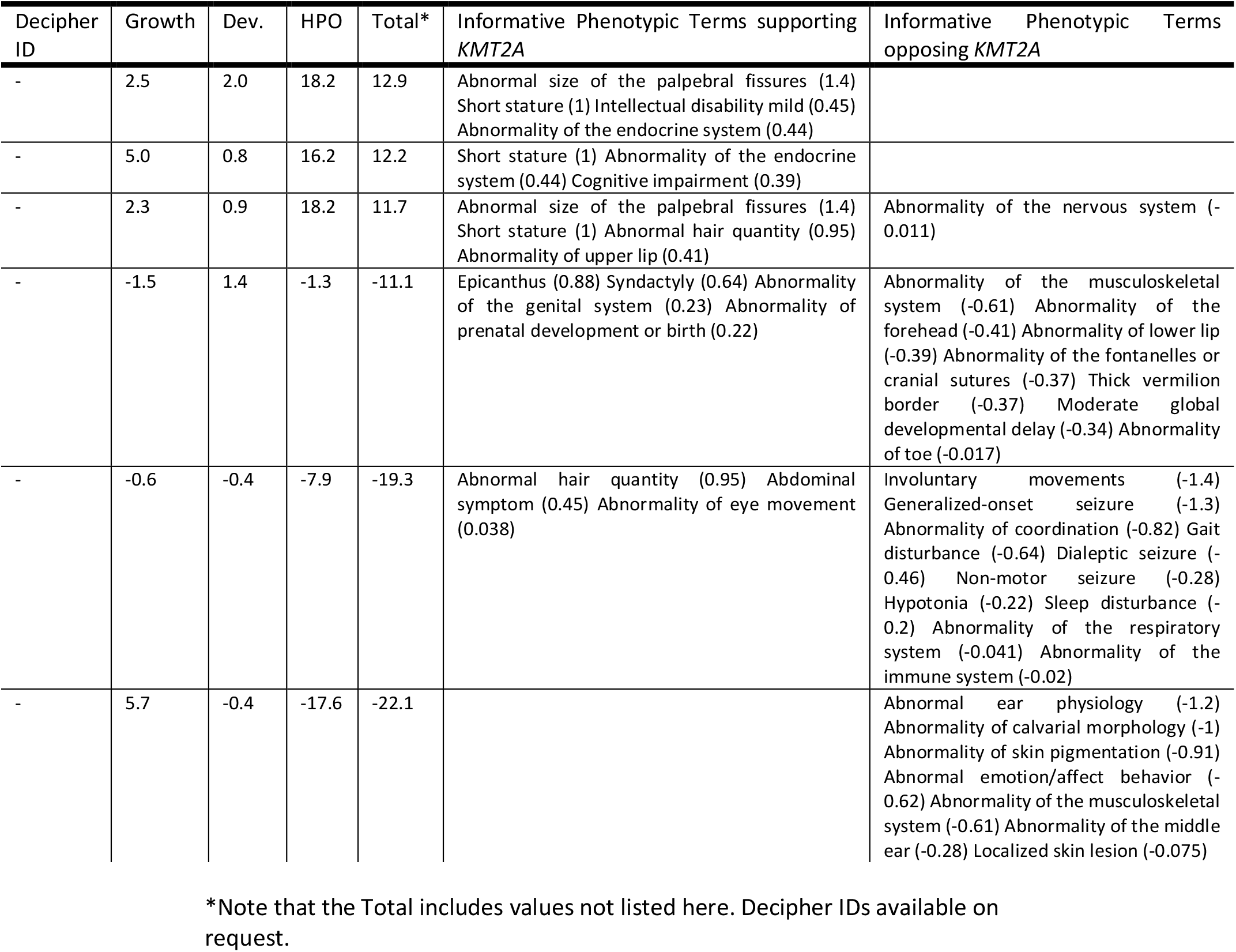
Clinical evidence supporting or opposing the diagnosis of *KMT2A* in probands with a pathogenic *de novo* variant.

As would be expected, gender alone was a poor predictor, however, this attribute has information by AUC for three genes: *SMC1A, MECP2* and *DDX3X* (**Figure 3B**). A strong female bias is known for these X-linked syndromes.

We next optimised the F1 score for each gene by combining the likelihood ratios for each data type using five weights *w*_*0*_*-w*_*4*_ (equation 2). For a given set of weights, a value from equation 2 greater than 0 is a classification to the gene, from which true and false positives can be determined and F1 calculated.

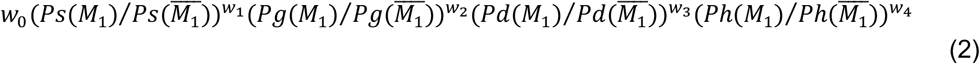

In (2), *p*(*D*|*M*) is abbreviated to *ps*(*M*) for gender, and similarly for growth (*Pg*), development (*Pd*), and HPO (*Ph*). Simulated annealing implemented in the R package GenSA was used to find the optimal weights (Web Resources).

For all genes, optimisation improved F1 over any individual data source (**Figure 3C**) achieving F1 scores greater than 0.7 for 12 genes when testing on training data. Genes with lower numbers of cases were high in the ranking, indicating that good models can be found for them, however, overfitting in the original model training may be at play. The F1 score was generally reduced in leave-one-out cross-validation where 14 genes had F1 0.3-0.5, namely, *ARID1B, CHD7, DYRK1A, EFTUD2, EP300, FOXP1, ITPR1, KIF1A, KMT2A, NSD1, PPP2R5D, PTEN PTPN11* and *SATB2* (**Figure 3D, S3**). Genes with larger numbers of cases tended to have higher F1 (Pearson correlation 0.27; p=0.015).

We also examined the distribution of likelihoods of a diagnosis **(Figure 3E**) versus all others **(Figure 3F**) from HPO gene models. This analysis highlighted a number of genes, including *KMT2A*, where the evidence from the fit to the HPO model did not outweigh the prior for many individuals and hence giving false negatives.

We then asked if an optimised HPO classifier would rival the combined data classifier. Optimal values were found for *w*_*0*_ and *w*_*4*_ (for the prior and HPO likelihood) using the same procedure. Using only HPO terms gave 147 fewer true positives (8.5% of the 1730 cases) and 2041 more false positives in total (summing false positives over 77 genes). Per gene, median recall was reduced from 0.57 to 0.5, and median precision from 0.64 to 0.21. The benefits of additional phenotypic information, specifically growth and development data, are clear from these results. As an additional comparison, classifiers based on top terms by TF IDF IC and by disease model were also assessed and found not perform as well as IPT-based classifiers (Supplemental Methods).

To further investigate the generality of each model in each data type, we assessed growth, development and HPO models through their contribution to the optimised log likelihood for all probands for each gene (**Figure 4A**). This revealed models that worked well in decision-making did not necessarily capture all diagnosed individuals, for example, growth in *NSD1* and development in *GRIN2B* captured distinctive subsets of probands. To visualise this across all 77 genes, we selected probands at quartiles 1, 2 and 3 representing poor, typical and good fits to the gene models (**Figure 4B**). Where the scaled values were negative, the model contradicted the assigned diagnosis. We found HPO models agreed with the correct diagnosis at each quartile. Defining models to generalise well as those in which a positive likelihood ratio is found for the median proband, all but four growth models generalised well. However, development models for 22 genes (29%) failed to generalise by this criterion.

**Figure 4.**
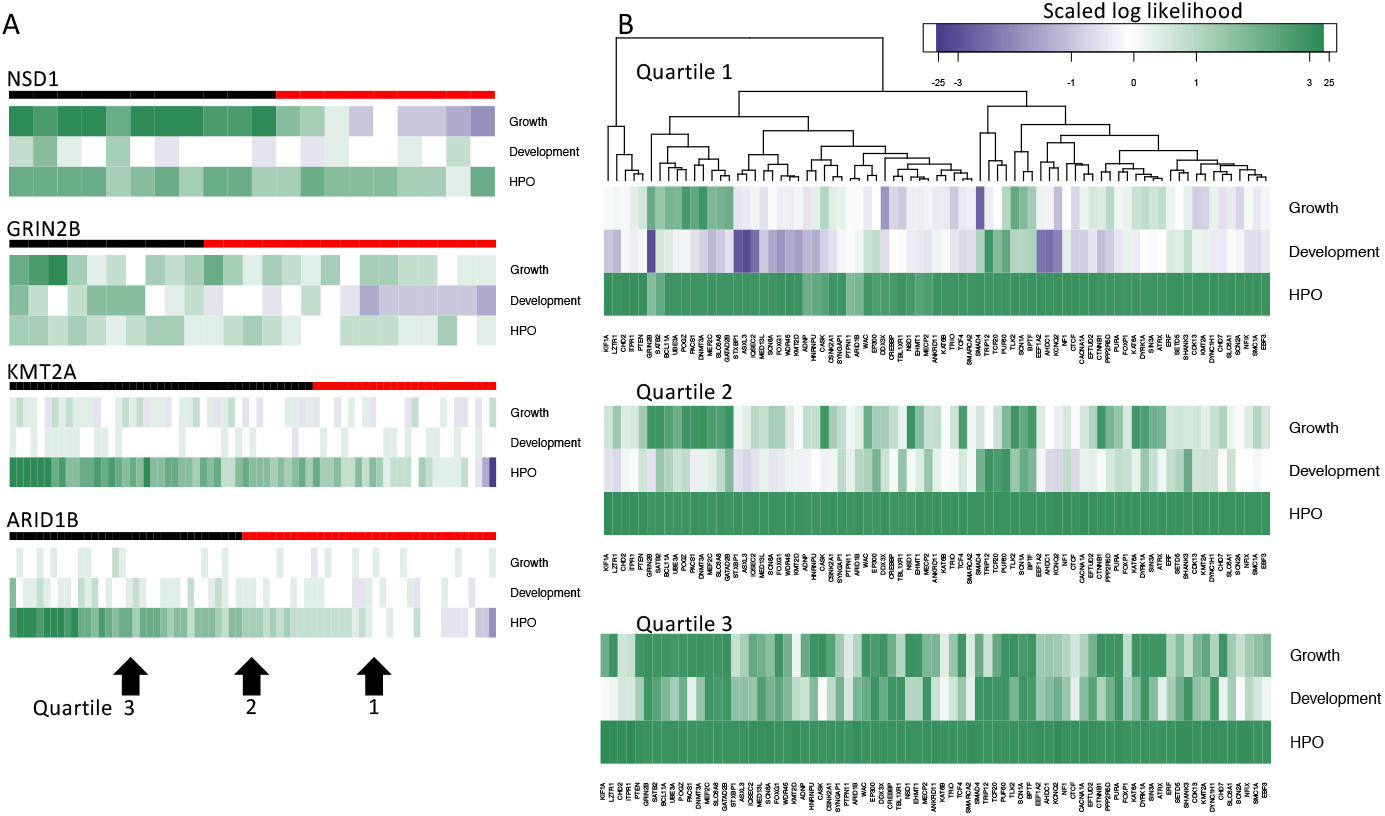
The contribution of data type to diagnosis varies by gene. (A) Heatmaps of log likelihood per proband (column) for each data type (row) for selected genes. Values are scaled by optimisation weight and columns ordered right to left from highest to lowest total likelihood (negative values in blue, positive in green). Upper bar shows true positives in black, false negatives in red. (B) Heatmaps of log likelihood multiplied by optimisation weight per gene (column) for each data type (row). Heatmaps show values for probands at quartiles 1, 2 and 3 successively for each data type. The hierarchical clustering reflects groupings at quartile 1. For example, for *GRIN2B*, the proband with the first quartile score fits the development model poorly (indicated in blue), the median *GRIN2B* proband has a small negative contribution from development and the contribution from development is positive by the third quartile. Only positive contributions are found from quartile 3, and HPO has a positive contribution at each quartile. The colour scale truncates absolute values above 3 in order to focus on the range -3 to 3.

In summary, we found that HPO terms are the best individual predictor of a correct diagnosis for most genes, however 17% (13/77 genes) were better predicted by growth or development, and prediction from combined data performed better than any individual source. Predictions from combined data also gave notably fewer false positives than prediction from HPO terms alone as median precision per increased to 0.64 from 0.21 when only HPO terms were used. While more cases are to be preferred when building models, we found gene models with good predictive performance on cross-validation could be built from a relatively small number of cases (n≥10). The derivation of likelihood ratios from proband annotations using HPO-encoded disease models of LIRICAL^6^ is closest to our approach. However, rather than define HPO models per gene, we began with the extensive database of proband annotations of the DDD study from which we are able to assess HPO term usage in clinical practice irrespective of diagnosis – our disease models are then computed from the observed annotations for each gene.

Bayesian methods are recognised as adding quantitative rigour to the combination of evidence for and against variant pathogenicity in rare disease, whilst making explicit any assumptions regarding strength of evidence and disease prevalence^27^. We extend the application of this paradigm to phenotypic data making use of the extensive acquisition of growth measures and developmental milestones in addition to HPO terms in the DDD study. This approach could be extended by the incorporation of additional phenotypic data (e.g., images, epigenomic profiles, biochemical assays, etc.) to further improve gene-disease models and make them more applicable to other rare disorders. Although phenotypic models are unlikely to be sufficiently predictive by themselves, particularly for genetically heterogenous disorders such as DD, they can be used to update posterior probabilities found from genomic analyses of variant pathogenicity^27^ and thus have an important role in increasing the robustness of a diagnosis.

Our findings emphasise the importance of systematically collecting and storing the quantitative patient data often recorded health records in computationally accessible formats to complement genomic analysis and clinical terminologies.

## Data Availability

Data are available at https://www.deciphergenomics.org/

## Acknowledgements

The DDD study presents independent research commissioned by the Health Innovation Challenge Fund [grant number HICF-1009-003], a parallel funding partnership between Wellcome and the Department of Health, and the Wellcome Sanger Institute [grant number WT098051]. The views expressed in this publication are those of the author(s) and not necessarily those of Wellcome or the Department of Health. The study has UK Research Ethics Committee approval (10/H0305/83, granted by the Cambridge South REC, and GEN/284/12 granted by the Republic of Ireland REC). The research team acknowledges the support of the National Institute for Health Research, through the Comprehensive Clinical Research Network. This study makes use of DECIPHER (https://www.deciphergenomics.org/), which is funded by the Wellcome. HVF is supported by the Wellcome Trust [award 200990/Z/16/Z] ‘Designing, developing and delivering integrated foundations for genomic medicine’. The research team acknowledges the support of the National Institute for Health Research, through the Comprehensive Clinical Research Network. DRF is funded as part of the MRC Human Genetics Unit grant to the University of Edinburgh. CAS and SA are supported by MRC Core funding to the MRC Human Genetics Unit (MRC grant MC_UU_00007/16).

## Web Resources

DECIPHER, https://deciphergenomics.org

genSA, https://CRAN.R-project.org/package=GenSA HPO, https://hpo.jax.org/app/

HPO, Gene Models http://purl.obolibrary.org/obo/hp/hpoa/genes_to_phenotype.txt IMPROVE-DD scripts, https://github.com/Stuart-Aitken/IMPROVE-DD naivebayes, https://CRAN.R-project.org/package=naivebayes

