## Supplemental Information for "IMPROVE-DD: Integrating Multiple Phenotype Resources Optimises Variant Evaluation in genetically determined Developmental Disorders"

### Methods

##### Classification and optimisation

Classifiers for nominal and continuous data were implemented using the naivebayes R package (Web Resources). For all growth and development data, the bandwidth of the nrd0 kernel to was increased to 1.5 to remove overfitting.

Optimisation was performed using the genSA R package (Web Resources). The inputs to optimisation were the likelihood ratios calculated by the models for each data source for all individuals. As the prior had been accounted for in all four cases, the priors were balanced to equality between the gene model and the alternative (as in **Figure 3E** and **3F**) and $w_0$ included in the optimisation as replacement. Optimisation maximised F1 by combining the likelihood values through the five weights as defined in equation (2).

A sensitivity analysis of parameter values around the optimal (replacing all parameters in each of 1000 iterations by sample from a normal distribution mean equal to the optimal, sd = 1/10 |optimal|) resulted in F1 values narrowly distributed around a reduced mean (**Figure S5**). The relatively small reduction in F1 indicates that non-optimal parameters cause the mis-classification of only a small number of cases.

##### Term selection and term probabilities

Annotations to HPO terms directly assigned to probands (4182 terms were used across 13439 individuals with a median of 6 terms per proband) were propagated to all parent terms, expanding the annotation to 5153 terms with a median of 40 terms per proband.

157 informative phenotypic terms were selected according to usage as decribed in the main text (**Figure 1D** and **1E**) with a median use of 10 terms per proband. Alternative thresholds for term inclusion were explored, and we concluded that the choice of thresholds was not critical.

Gene models were defined by the probabilities of the 157 IPTs in the annotations to diagnosed individuals *D*. The probability of each *IPT* in *D* was given by the m-estimate:

$$P(IPT) = {A(IPT,D) + 1} \Big/ {(\sum_{t=1}^{t=157} A(t,D)) + 100}$$

where the term usage $A(t,D)$ was calculated from the modified annotation matrix as the sum over *t* (column in matrix) for probands (rows) assigned to *D*. The denominator normalises the counts to a specific term by the total annotations made to the 157 IPTs plus a number representing the vocabulary size.

When computing the probability of a set of IPT annotations in a case $C_i$ for a given gene model, the product of probabilities was scaled to adjust for the number of IPTs used in this case. Where $C_i$ had $m$ IPTs the geometric mean probability was raised to the power of 10 (the median number of IPTs per case):

$$P(C_i\ GeneModel) = \prod_{j=1}^{m} P(IPTj)^{10/m}$$

This scaling made the resulting probabilities comparable across probands.

To investigate the impact of alternative term selection strategies, classifiers based on terms selected by TF IDF IC, or from HPO disease models were evaluated. In both cases, the 50 most informative terms per gene were selected (disease model terms were ordered by TF IDF IC) and term probability was based on term frequency in disease with Laplace smoothing:

$$P(HPO) = {A(HPO, D) + 1}\Big/{(|D| + 2)}$$

AUC and F1 were typically lower in gene-specific classifiers than for IPT classifiers when testing on training data (**Figure S4**). We found the occurrence of an HPO term of the gene model in a case to essentially guarantee classification to that model: Mean recall was high (0.79) showing that DDD cases indeed matched these disease models but mean precision was low (0.03). Even though we selected terms with the highest IC per gene, they occurred in other diagnoses giving many false positives.

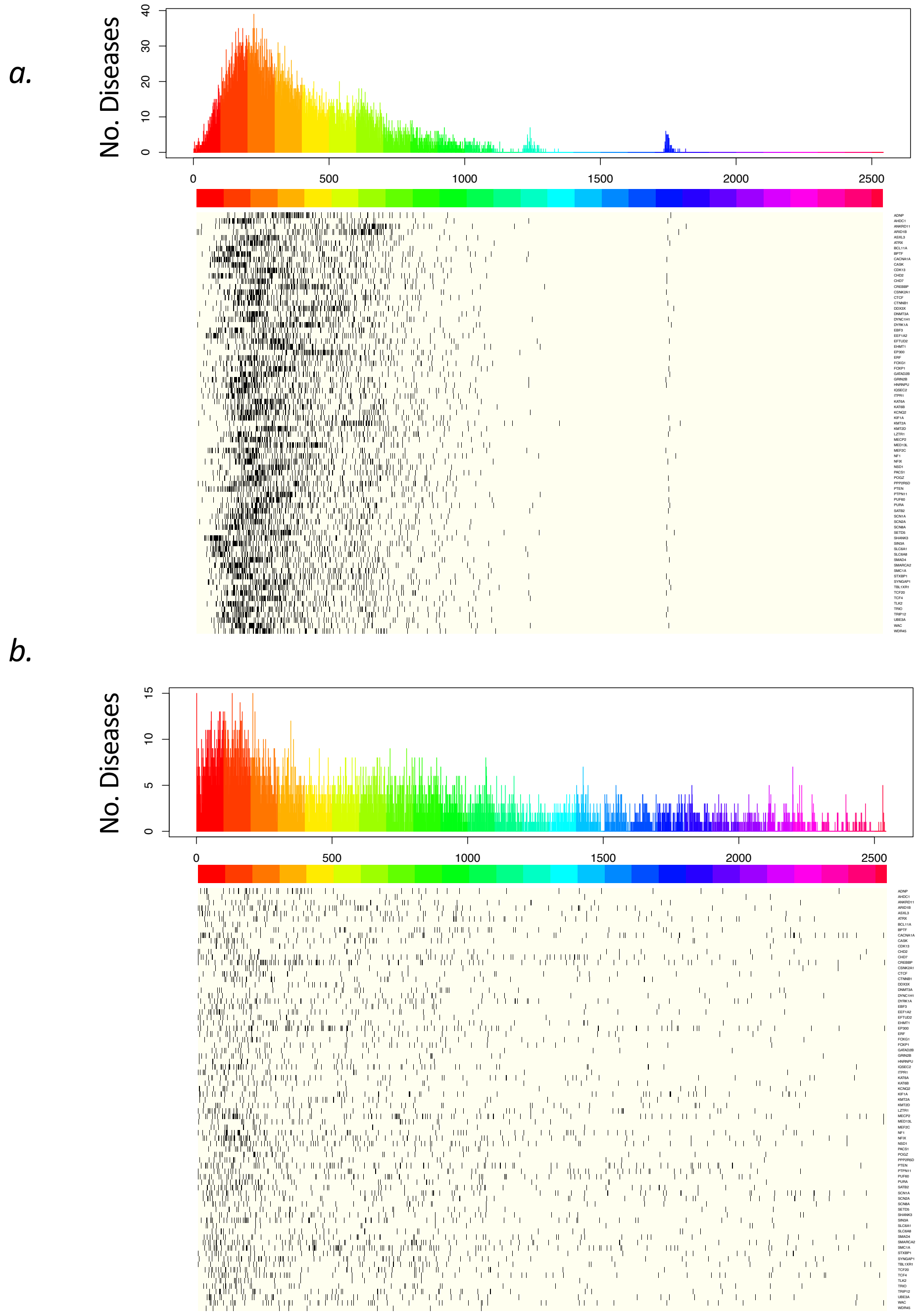


**Figure S1. Disease model term ranking**

a) Heatmap showing for each gene (row) the occurrence of an informative phenotypic term (IPT) in a ranking of all HPO terms by TF IDF IC. HPO terms are ordered left to right by decreasing TF IDF IC. Top panel shows the number of diseases for which a disease model term is found in rank i (from 1 to 2500), colours indicate scale, each covers 100 positions. Row order is alphabetical by gene name.

b) Heatmap showing for each gene (row) the occurrence of an term in the disease model for that gene in a ranking of all HPO terms by TF IDF IC.

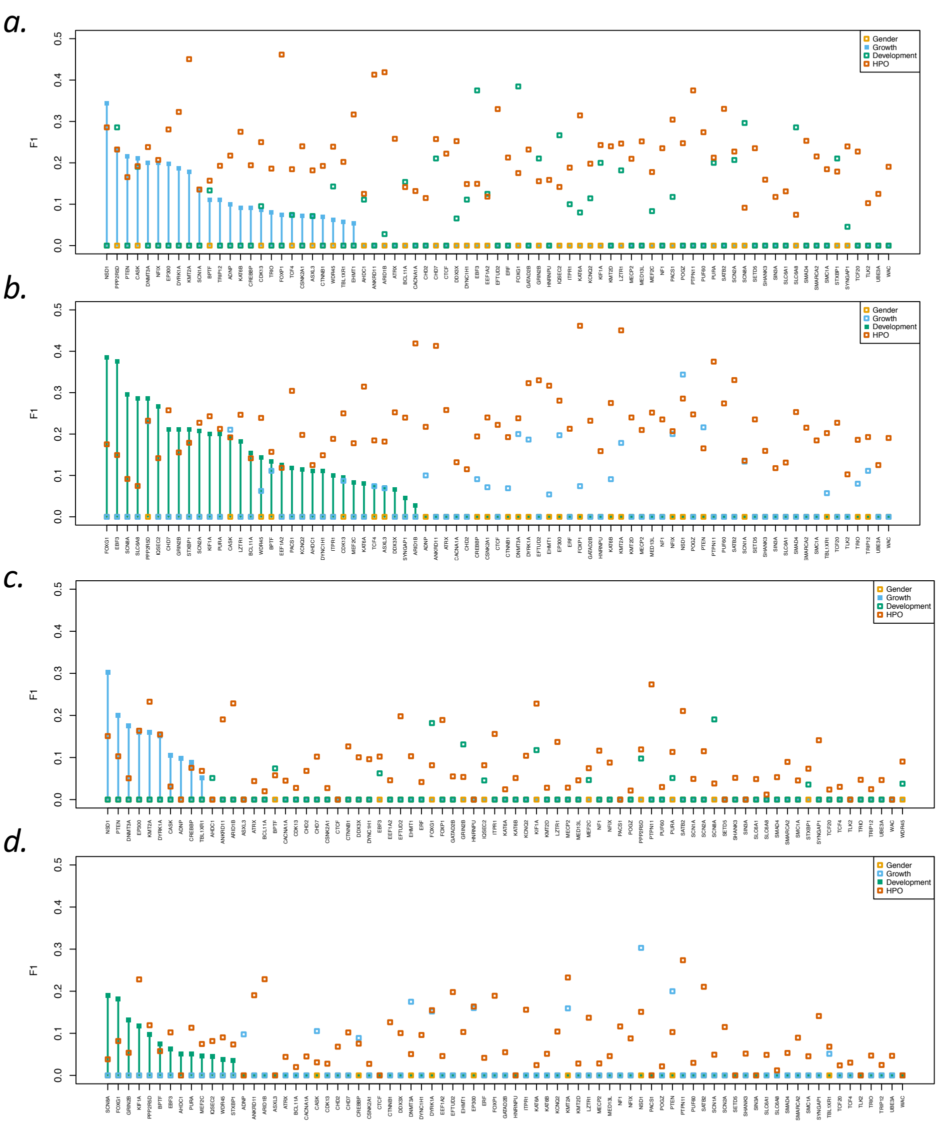


**Figure S2. Classifier performance.**

a) F1 per gene, testing on training data, ordered by the performance in growth (highlighted by the vertical bars and filled symbols) and ordered by development b).

c) F1 per gene, from a leave-one-out cross-validation, ordered by the performance in growth, and ordered by development d).

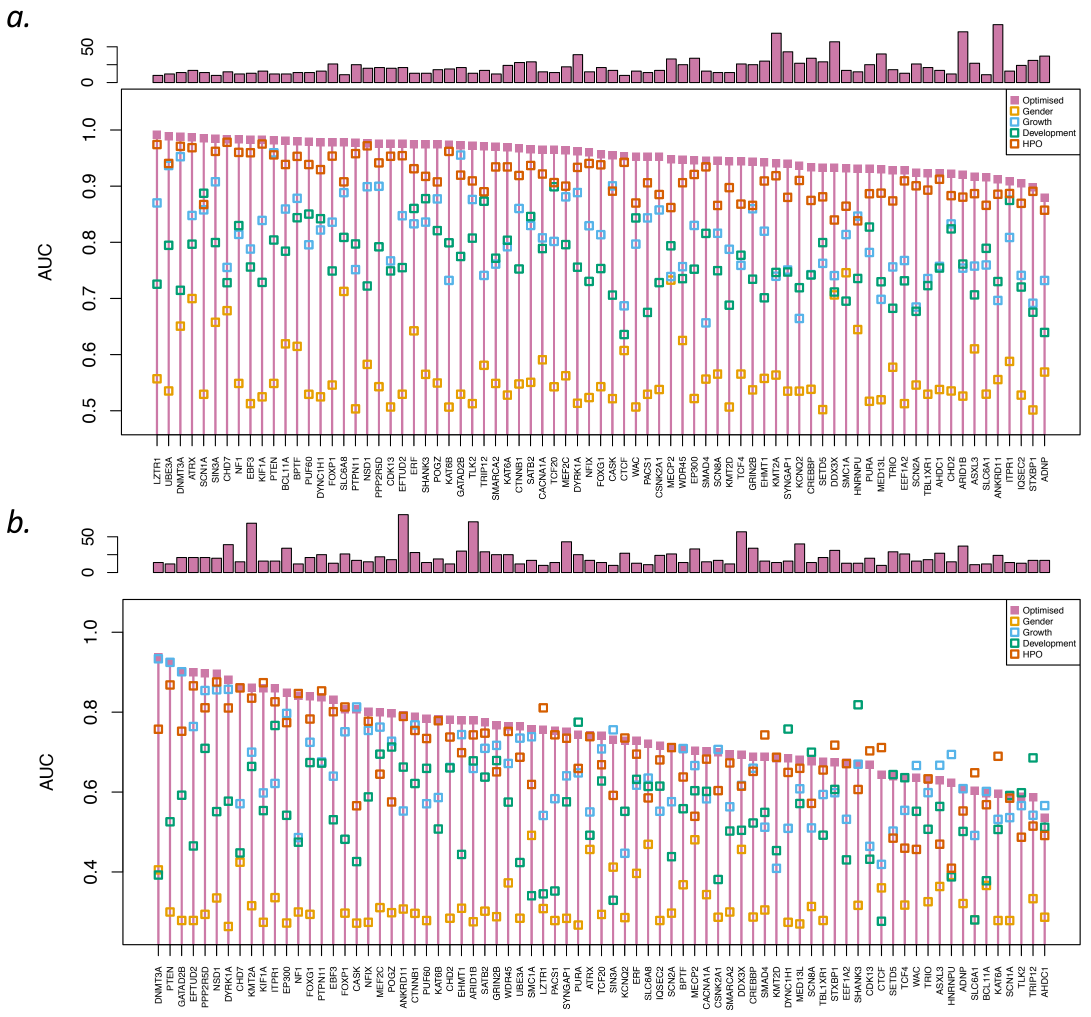


**Figure S3. AUC on optimisation.**

a) AUC from all data types when testing on training data is shown by symbols, closed symbols and vertical lines highlight the AUC after optimising for F1.

b) AUC from all data types in cross-validation is shown by symbols, closed symbols and vertical lines highlight the AUC after optimising for F1.

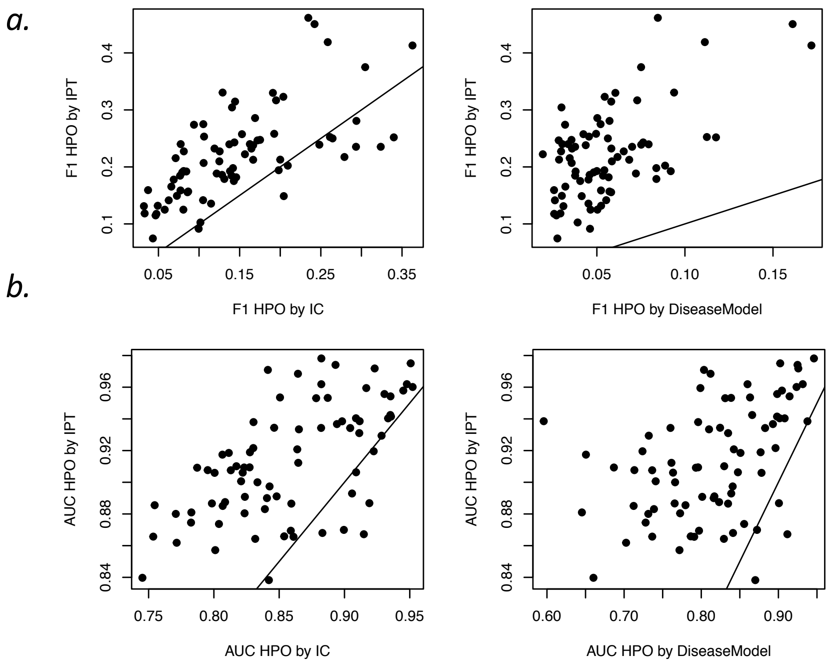


**Figure S4. Comparison between IPT and alternative term sets for classification.**

a) Scatterplot of F1 from IPTs against F1 from the top 50 terms per gene selected by TF IDF IC (left), symbols are genes, and against the top 50 disease model terms per gene selected by TF IDF IC.

b) Scatterplot of AUC from IPTs against AUC from the top 50 terms per gene selected by TF IDF IC (left) and against the top 50 disease model terms per gene selected by TF IDF IC.

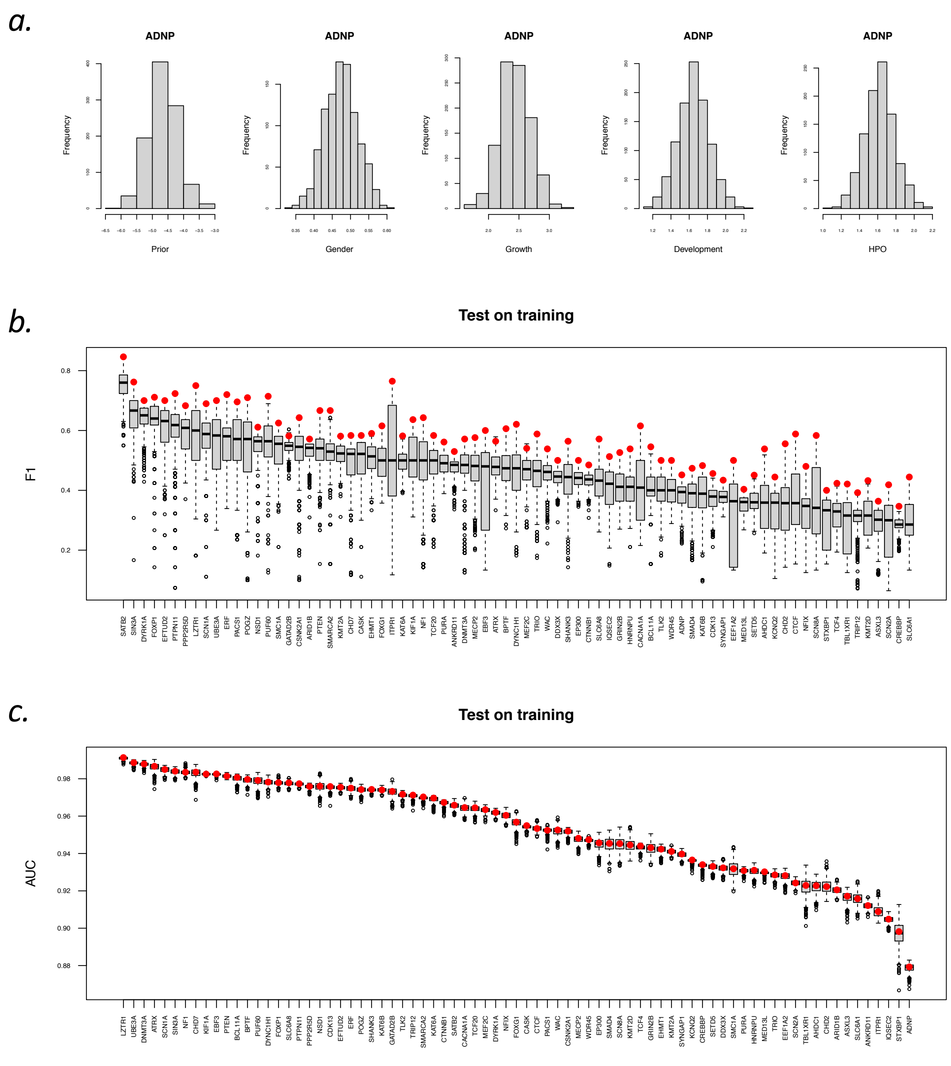


**Figure S5. Sensitivity analysis.**

a) Histograms of parameter values used in the sensitivity analysis of ADNP.

b) Boxplot of F1 per gene from 1000 random samples of the optimisation weights. F1 from optimal weights in red.

c) Boxplot of AUC per gene from 1000 random samples of the optimisation weights. AUC from optimal weights in red.

| | | | | | | | | |
|---|---|---|---|---|---|---|---|---|
| topleve | HP:0000707 | Abnormality of the nervous system | toplevel: | HP:0000598 | Abnormality of the ear | topleve | HP:0033127 | Abnormality of the musculoskeletal system |
| child: | HP:0000717 | Autism | child: | HP:0000357 | Abnormal location of ears | child: | HP:0000765 | Abnormal thorax morphology |
| | HP:0000736 | Short attention span | | HP:0000370 | Abnormality of the middle ear | | HP:0000925 | Abnormality of the vertebral column |
| | HP:0000752 | Hyperactivity | | HP:0000377 | Abnormality of the pinna | | HP:0001252 | Hypotonia |
| | HP:0001256 | Intellectual disability, mild | | HP:0031704 | Abnormal ear physiology | | HP:0001276 | Hypertonia |
| | HP:0001270 | Motor delay | toplevel: | HP:0025354 | Abnormal cellular phenotype | | HP:0001367 | Abnormal joint morphology |
| | HP:0001288 | Gait disturbance | [no child terms] | | | | HP:0001371 | Flexion contracture |
| | HP:0001328 | Specific learning disability | toplevel: | HP:0000478 | Abnormality of the eye | | HP:0001382 | Joint hypermobility |
| | HP:0001344 | Absent speech | child: | HP:0000316 | Hypertelorism | | HP:0001388 | Joint laxity |
| | HP:0002118 | Abnormal cerebral ventricle morpholog | | HP:0000490 | Deeply set eye | | HP:0003549 | Abnormality of connective tissue |
| | HP:0002197 | Generalized-onset seizure | | HP:0000496 | Abnormality of eye movement | | HP:0009115 | Aplasia/hypoplasia involving the skeleton |
| | HP:0002342 | Intellectual disability, moderate | | HP:0000504 | Abnormality of vision | | HP:0009122 | Aplasia/hypoplasia affecting bones of the axi |
| | HP:0002360 | Sleep disturbance | | HP:0000508 | Ptosis | | HP:0011314 | Abnormality of long bone morphology |
| | HP:0002493 | Upper motor neuron dysfunction | | HP:0000539 | Abnormality of refraction | | HP:0011805 | Abnormal skeletal muscle morphology |
| | HP:0004305 | Involuntary movements | | HP:0000553 | Abnormal uvea morphology | | HP:0030084 | Clinodactyly |
| | HP:0007370 | Aplasia/Hypoplasia of the corpus callosu | | HP:0004328 | Abnormal anterior eye segment morphology | | HP:0100261 | Abnormal tendon morphology |
| | HP:0010864 | Intellectual disability, severe | | HP:0004329 | Abnormal posterior eye segment morphology | topleve | HP:0001871 | Abnormality of blood and blood-forming tissu |
| | HP:0010993 | Abnormality of the cerebral subcortex | | HP:0008056 | Aplasia/Hypoplasia affecting the eye | [no child terms] | | |
| | HP:0011146 | Dialeptic seizure | toplevel: | HP:0001197 | Abnormality of prenatal development or birtl | topleve | HP:0001574 | Abnormality of the integument |
| | HP:0011282 | Abnormality of hindbrain morphology | [no child terms] | | | child: | HP:0000499 | Abnormal eyelash morphology |
| | HP:0011342 | Mild global developmental delay | toplevel: | HP:0000152 | Abnormality of head or neck | | HP:0001000 | Abnormality of skin pigmentation |
| | HP:0011343 | Moderate global developmental delay | toplevel: | HP:0000598 | Abnormality of the ear | | HP:0001597 | Abnormality of the nail |
| | HP:0011344 | Severe global developmental delay | child: | HP:0000357 | Abnormal location of ears | | HP:0010720 | Abnormal hair pattern |
| | HP:0011443 | Abnormality of coordination | | HP:0000370 | Abnormality of the middle ear | | HP:0011122 | Abnormality of skin physiology |
| | HP:0012447 | Abnormal myelination | | HP:0000377 | Abnormality of the pinna | | HP:0011354 | Generalized abnormality of skin |
| | HP:0020219 | Motor seizure | | HP:0031704 | Abnormal ear physiology | | HP:0011355 | Localized skin lesion |
| | HP:0033259 | Non-motor seizure | toplevel: | HP:0025354 | Abnormal cellular phenotype | | HP:0011356 | Regional abnormality of skin |
| | HP:0100543 | Cognitive impairment | [no child terms] | | | | HP:0011362 | Abnormal hair quantity |
| | HP:0100851 | Abnormal emotion/affect behavior | toplevel: | HP:0000478 | Abnormality of the eye | | HP:0100037 | Abnormality of the scalp hair |
| topleve | HP:0001939 | Abnormality of metabolism/homeostasis | child: | HP:0000316 | Hypertelorism | topleve | HP:0040064 | Abnormality of limbs |
| [no child terms] | | | | HP:0000490 | Deeply set eye | child: | HP:0001159 | Syndactyly |
| topleve | HP:0000818 | Abnormality of the endocrine system | | HP:0000496 | Abnormality of eye movement | | HP:0001172 | Abnormal thumb morphology |
| [no child terms] | | | | HP:0000504 | Abnormality of vision | | HP:0001211 | Abnormal fingertip morphology |
| topleve | HP:0000769 | Abnormality of the breast | | HP:0000508 | Ptosis | | HP:0001763 | Pes planus |
| [no child terms] | | | | HP:0000539 | Abnormality of refraction | | HP:0001780 | Abnormality of toe |
| topleve | HP:0001507 | Growth abnormality | | HP:0000553 | Abnormal uvea morphology | | HP:0004097 | Deviation of finger |
| child: | HP:0000098 | Tall stature | | HP:0004328 | Abnormal anterior eye segment morphology | | HP:0004207 | Abnormal 5th finger morphology |
| | HP:0001511 | Intrauterine growth retardation | | HP:0004329 | Abnormal posterior eye segment morphology | | HP:0005656 | Positional foot deformity |
| | HP:0004322 | Short stature | | HP:0008056 | Aplasia/Hypoplasia affecting the eye | | HP:0005918 | Abnormal finger phalanx morphology |
| | HP:0004323 | Abnormality of body weight | toplevel: | HP:0001197 | Abnormality of prenatal development or birtl | | HP:0005922 | Abnormal hand morphology |
| topleve | HP:0001608 | Abnormality of the voice | [no child terms] | | | | HP:0005927 | Aplasia/hypoplasia involving bones of the ha |
| [no child terms] | | | toplevel: | HP:0000152 | Abnormality of head or neck | | HP:0006494 | Aplasia/Hypoplasia involving bones of the fe |
| topleve | HP:0002715 | Abnormality of the immune system | toplevel: | HP:0000598 | Abnormality of the ear | | HP:0006496 | Aplasia/hypoplasia involving bones of the up |
| [no child terms] | | | child: | HP:0000357 | Abnormal location of ears | | HP:0009484 | Deviation of the hand or of fingers of the hai |
| topleve | HP:0001626 | Abnormality of the cardiovascular syste | | HP:0000370 | Abnormality of the middle ear | | HP:0009810 | Abnormality of upper limb joint |
| child: | HP:0001627 | Abnormal heart morphology | | HP:0000377 | Abnormality of the pinna | | HP:0009815 | Aplasia/hypoplasia of the extremities |
| | HP:0002597 | Abnormality of the vasculature | | HP:0031704 | Abnormal ear physiology | | HP:0011927 | Short digit |
| | HP:0011025 | Abnormal cardiovascular system physiol | toplevel: | HP:0025354 | Abnormal cellular phenotype | | HP:0040069 | Abnormal lower limb bone morphology |
| | HP:0025015 | Abnormal vascular morphology | [no child terms] | | | | HP:0040070 | Abnormal upper limb bone morphology |
| topleve | HP:0000119 | Abnormality of the genitourinary systen | toplevel: | HP:0000478 | Abnormality of the eye | | HP:0045060 | Aplasia/hypoplasia involving bones of the ex |
| child: | HP:0000078 | Abnormality of the genital system | child: | HP:0000316 | Hypertelorism | | HP:0100491 | Abnormality of lower limb joint |
| | HP:0000079 | Abnormality of the urinary system | | | | | HP:0100807 | Long fingers |
| topleve | HP:0002664 | Neoplasm | | | | | HP:0100871 | Abnormality of the palm |
| [no child terms] | | | | | | topleve | HP:0002086 | Abnormality of the respiratory system |
| | | | | | | [no child terms] | | |
| | | | | | | topleve | HP:0025142 | Constitutional symptom |
| | | | | | | [no child terms] | | |
| | | | | | | topleve | HP:0025031 | Abnormality of the digestive system |
| | | | | | | child: | HP:0004298 | Abnormality of the abdominal wall |
| | | | | | | | HP:0011024 | Abnormality of the gastrointestinal tract |
| | | | | | | | HP:0011458 | Abdominal symptom |
| | | | | | | | HP:0012719 | Functional abnormality of the gastrointestina |
| | | | | | | | HP:0025033 | Abnormality of digestive system morphology |

**Table S1. Informative Phenotypic Terms.**
